# Eight-Fold Increased COVID-19 Mortality in Autosomal Dominant Tubulointerstitial Kidney Disease due to *MUC1* Mutations: An Observational Study

**DOI:** 10.1101/2024.07.03.24309887

**Authors:** Kendrah O. Kidd, Adrienne H. Williams, Abbigail Taylor, Lauren Martin, Victoria Robins, John A. Sayer, Eric Olinger, Holly R. Mabillard, Gregory Papagregoriou, Constantinos Deltas, Christoforos Stavrou, Peter J. Conlon, Richard Edmund Hogan, Elhussein A.E. Elhassan, Drahomíra Springer, Tomáš Zima, Claudia Izzi, Alena Vrbacká, Lenka Piherová, Michal Pohludka, Martin Radina, Petr Vylet’al, Katerina Hodanova, Martina Zivna, Stanislav Kmoch, Anthony J. Bleyer

**Author notes:** CORRESPONDING AUTHOR Anthony J. Bleyer, Sr., M.D., M.S. Section on Nephrology, Wake Forest University School of Medicine Medical Center Blvd. Winston-Salem, NC 27157.

## Abstract

**Background:** *MUC1* and *UMOD* pathogenic variants cause autosomal dominant tubulointerstitial kidney disease (ADTKD). *MUC1* is expressed in kidney, nasal mucosa and respiratory tract, while *UMOD* is expressed only in kidney. Due to haplo-insufficiency ADTKD-*MUC1* patients produce approximately 50% of normal mucin-1.

**Methods:** To determine whether decreased mucin-1 production was associated with an increased COVID-19 risk, we sent a survey to members of an ADTKD registry in September 2021, after the initial, severe wave of COVID-19. We linked results to previously obtained ADTKD genotype and plasma CA15-3 (mucin-1) levels and created a longitudinal registry of COVID-19 related deaths.

**Results:** Surveys were emailed to 637 individuals, with responses from 89 ADTKD-*MUC1* and 132 ADTKD-*UMOD* individuals. 19/83 (23%) ADTKD-*MUC1* survey respondents reported a prior COVID-19 infection vs. 14/125 (11%) ADTKD-*UMOD* respondents (odds ratio (OR) 2.35 (95%CI 1.60-3.11, *P* = 0.0260). Including additional familial cases reported from survey respondents, 10/41 (24%) ADTKD-*MUC1* individuals died of COVID-19 vs. 1/30 (3%) with ADTKD-*UMOD*, with OR 9.21 (95%CI 1.22-69.32), *P* = 0.03. The mean plasma mucin-1 level prior to infection in 14 infected and 27 uninfected ADTKD-*MUC1* individuals was 7.06±4.12 vs. 10.21±4.02 U/mL (*P* = 0.035). Over three years duration, our longitudinal registry identified 19 COVID-19 deaths in 360 ADTKD-*MUC1* individuals (5%) vs. 3 deaths in 478 ADTKD-*UMOD* individuals (0.6%) (*P* = 0.0007). Multivariate logistic regression revealed the following odds ratios (95% confidence interval) for COVID-19 deaths: ADTKD-*MUC1* 8.4 (2.9-29.5), kidney transplant 5.5 (1.6-9.1), body mass index (kg/m^2^) 1.1 (1.0-1.2), age (y) 1.04 (1.0-1.1).

**Conclusions:** Individuals with ADTKD-*MUC1* are at an eight-fold increased risk of COVID-19 mortality vs. ADTKD-*UMOD* individuals. Haplo-insufficient production of mucin-1 may be responsible.

## INTRODUCTION

Mucin-1 is a membrane-anchored glycoprotein that provides physical protection to the epithelial surface of many tissues^1^ as well as performing other functions, including preventing infection,^2^ mediating inflammation,^3^ and modulating apoptosis.^4^ In autosomal dominant tubulointerstitial kidney disease (ADTKD) due to heterozygous frameshift variants in the *MUC1* gene (ADTKD-*MUC1*) (OMIM 174000),^5^ the wild-type allele synthesizes normal mucin-1, and the allele with the pathogenic *MUC1* variant produces a frameshift mucin-1 protein that deposits in the endoplasmic reticulum Golgi intermediate compartment (ERGIC).^6^ While abnormal protein deposition occurs within all epithelial cells expressing mucin-1^7^, clinical sequelae of the disease were thought to be limited to the kidney, with no respiratory, gastrointestinal, or skin manifestations previously identified.^8,9^ Affected individuals develop slowly progressive chronic kidney disease (CKD) that leads to kidney failure at a mean age of 45 years.^8^ ADTKD-*UMOD* (OMIM 162000, 603860) is phenotypically similar and often clinically indistinguishable from ADTKD-*MUC1* (OMIM 17400).^10^ Unlike mucin-1, uromodulin is expressed exclusively in the thick ascending limb of Henle.^11^ The median age of kidney failure in ADTKD-*UMOD* is 49 years.^12^ Both groups of individuals have excellent outcomes with kidney transplantation^13^, as the diseases do not recur in the transplanted kidneys and individuals usually have few other comorbid conditions.

Mucin-1 has been implicated as a protective factor in COVID-19, though this relationship has not been firmly established in human studies. Mucin-1 is produced in both the nares and the lungs, primary sites of COVID-19 infection.^14,15^ Mucin-1 in breast milk has been shown to inhibit SARS-CoV-2 infection^16^. In *in vitro* studies, mucin-1 was highly expressed on the surface of ACE2-positive respiratory epithelial cells, and the glycosylated extracellular domain of mucin-1 restricted SARS-CoV-2 binding and entry^17^.

Patients with ADTKD-*MUC1* may be at increased risk of COVID-19 due to haploinsufficiency of mucin-1. In ADTKD-MUC1, there is one wild-type allele that produces mucin-1 and one mutated allele that produces a truncated version of mucin-1 that deposits intracellularly and does not function as a normal mucin-1 protein. In a recent investigation, the mean plasma CA15-3 level, which measures mucin-1 levels with an immunoassay using the DF3 antibody,^18^ was 8.6 ± 4.3 U/mL in individuals with ADTKD-*MUC1* vs. 14.6 ± 5.6 U/mL in controls (*P* < 0.001).^19^ The mucin-1 content of plasma is derived primarily from the lungs.^20^ Thus, ADTKD-MUC1 patients produce less mucin-1, which may protect against COVID-19.

To determine if individuals with ADTKD-*MUC1* were at increased risk of COVID-19, we decided to compare the rates of COVID-19 infections between patients with ADTKD-*MUC1* and patients with ADTKD-*UMOD* in the Wake Forest Rare Inherited Kidney Disease registry. The registry attempts to recruit all individuals with ADTKD-*UMOD* and ADTKD-*MUC1* and includes 360 adults from 119 families affected with ADTKD-*MUC1* and 478 individuals from 171 families affected with ADTKD-*UMOD*. The registry has grown over the last two decades, with approximately 25% of individuals making contact with the Wake Forest Rare Inherited Kidney Disease Team independently without physician referral.^21^ The registry keeps in contact with family members through webinars, in-person meetings, email, and a private Facebook^©^ support page. Thus, there is a large registry containing individuals with ADTKD-*MUC1* and ADTKD-*UMOD*, without any perceived bias in interactions based on ADTKD type. The two diseases have a bland urinary sediment, slowly progressive chronic kidney disease leading to kidney failure at a mean age of approximately 45 years, and the absence of non-renal clinical manifestations.

To determine if individuals with ADTKD-*MUC1* were at increased risk of COVID-19, we first conducted a systematic survey of individuals with these two conditions in September 2021 (see **Figure 1**), with a specific goal of avoiding any ascertainment bias between the ADTKD-*UMOD* and ADTKD-*MUC1* groups. At that time, most individuals had had the opportunity to obtain COVID-19 vaccination, and the worst wave of COVID-19 deaths in the US had occurred. We informed patients in our registry of our interest in COVID-19 deaths in ADTKD-*UMOD* and ADKTD-*MUC1* patients and began tracking these occurrences.

**Figure 1.**
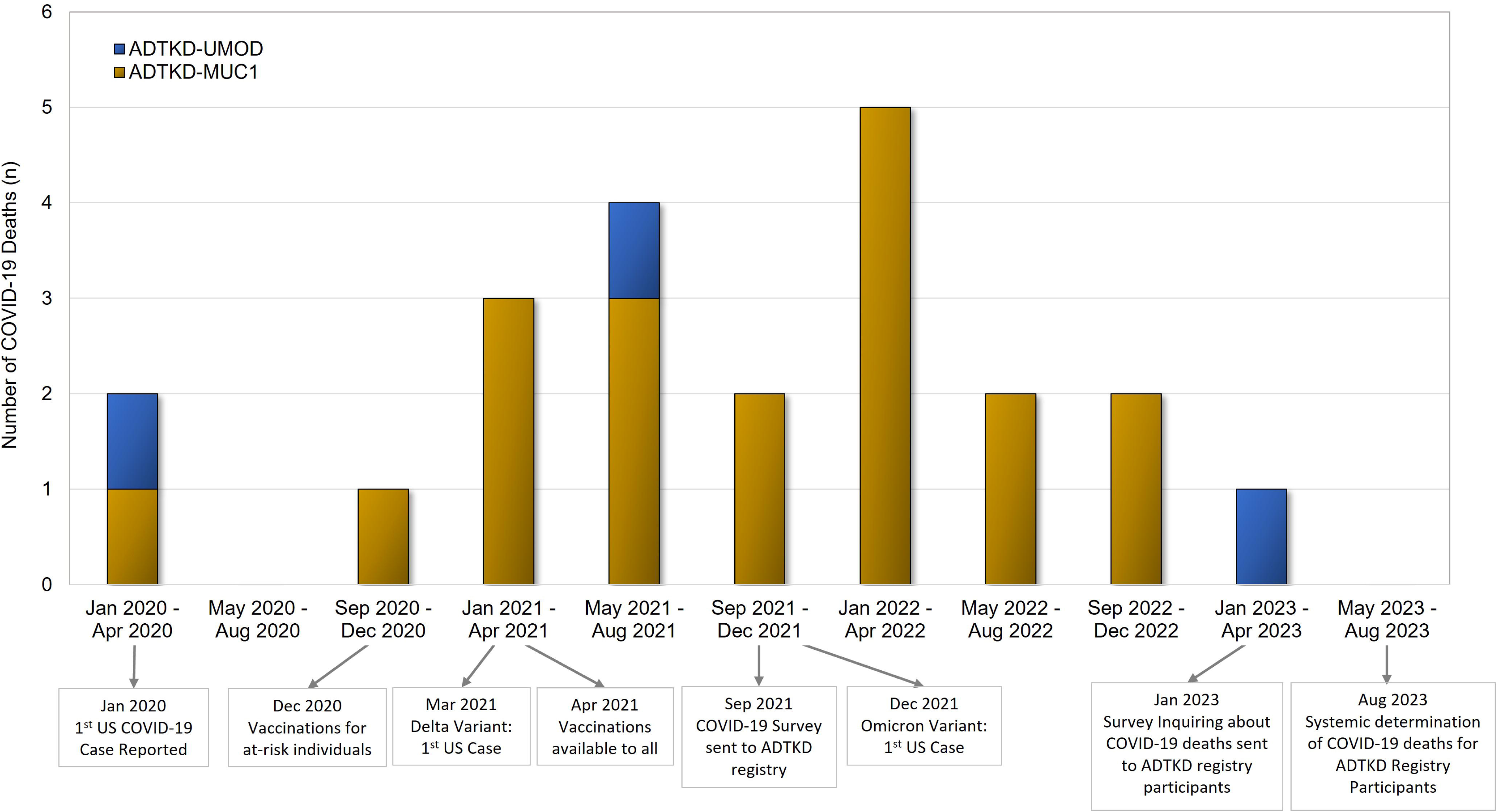
Study timeline and COVID-19 related deaths in ADTKD patients.

## MATERIALS AND METHODS

This study was approved by the Wake Forest Institutional Review Board (IRB00000352, Sub-study: COVID-19 Effects in Inherited Kidney Disease, approved September 8, 2021). We developed a survey and distributed it electronically to all individuals with ADTKD within our registry (see **Figure 1** and **Supplementary Material**). Survey data were collected between September 24, 2021 and November 1, 2021 and managed using REDCap electronic data capture tools hosted at Wake Forest School of Medicine. REDCap is a secure, web-based, National Institutes of Health (NIH)–sponsored application^22^ that supports confidential data capture for research studies. Information collected included: COVID-19 vaccination type and number of vaccines administered; COVID-19 infection characteristics including the date of first clinical symptoms, need for hospitalization, need for intensive care unit admission, and death or recovery from infection. Survey respondents provided information about their personal COVID-19 infections as well as those of family members, as family members may have died or become too sick to respond to our survey. For reported COVID-19 deaths, we spoke personally with family members to ascertain that the death was related to COVID-19. Data from the survey respondents were linked to other data in our registry that had been obtained prior to COVID-19 infections. This data included the ADTKD mutation (*UMOD* vs. *MUC1*) and plasma CA15-3 (mucin-1) levels. CA15-3 levels were measured as previously reported.^19^

We continued to ascertain COVID-19 deaths in our survey. In January 2023, an additional survey was sent to individuals with ADTKD that requested information about COVID-19 deaths. In August 2023, we reviewed all family trees of ADTKD-*UMOD* and ADTKD-*MUC1* families to identify new deaths by searching each name on-line for potential obituaries and then contacting family members about causes of death. We then performed a case-control study with cases including adults affected with ADTKD-*UMOD* and ADTKD-*MUC1* who died of COVID-19 and controls including adults affected with ADTKD-*UMOD* and ADTKD-*MUC1* who did not die from COVID-19.

### Statistical analysis

Survey questions were summarized with the addition of descriptive statistics and additional descriptive data were added from registry data when available. Associations were assessed using a generalized estimating equation to account for the family structure. We assumed exchangeable correlation within family and computed a robust sandwich estimator of variance.^23^ A compound-symmetry covariance structure was used to account for the correlation between family members for continuous outcomes.

### Genotyping

Individuals were genotyped for *UMOD* pathogenic variants using standard genetic techniques.^12^ For *MUC1* sequencing, a CLIA-approved mass spectrometry-based assay was performed the Broad Institute of MIT and Harvard, Cambridge, MA ^24^ or Illumina and/or PacBio® sequencing of *MUC1* PCR amplicon was performed by the Kmoch laboratory, First Faculty of Medicine, Prague, Czech Republic.^7^

## RESULTS

The initial survey was emailed to 637 individuals (see **Figure 2a**), including 256 patients with ADTKD-*MUC1* and 381 patients with ADTKD-*UMOD*. There were 257 respondents (35% response rate). The response rate for individuals with ADTKD-*MUC1* was 89/256(35%), and the response rate for individuals with ADTKD-*UMOD* was 132/381(35%). Compared to non-respondents, respondents were more likely to be White (97% vs. 90%, *P* = 0.02) and older (50.2±15.2 years vs. 47.0±16.0 years, *P* = 0.02) (see **Supplementary Table S1**). Of the respondents, six individuals with ADTKD-*MUC1* and seven with ADTKD-*UMOD* did not complete the survey about personal COVID-19 infection and were removed.

**Figure 2.**
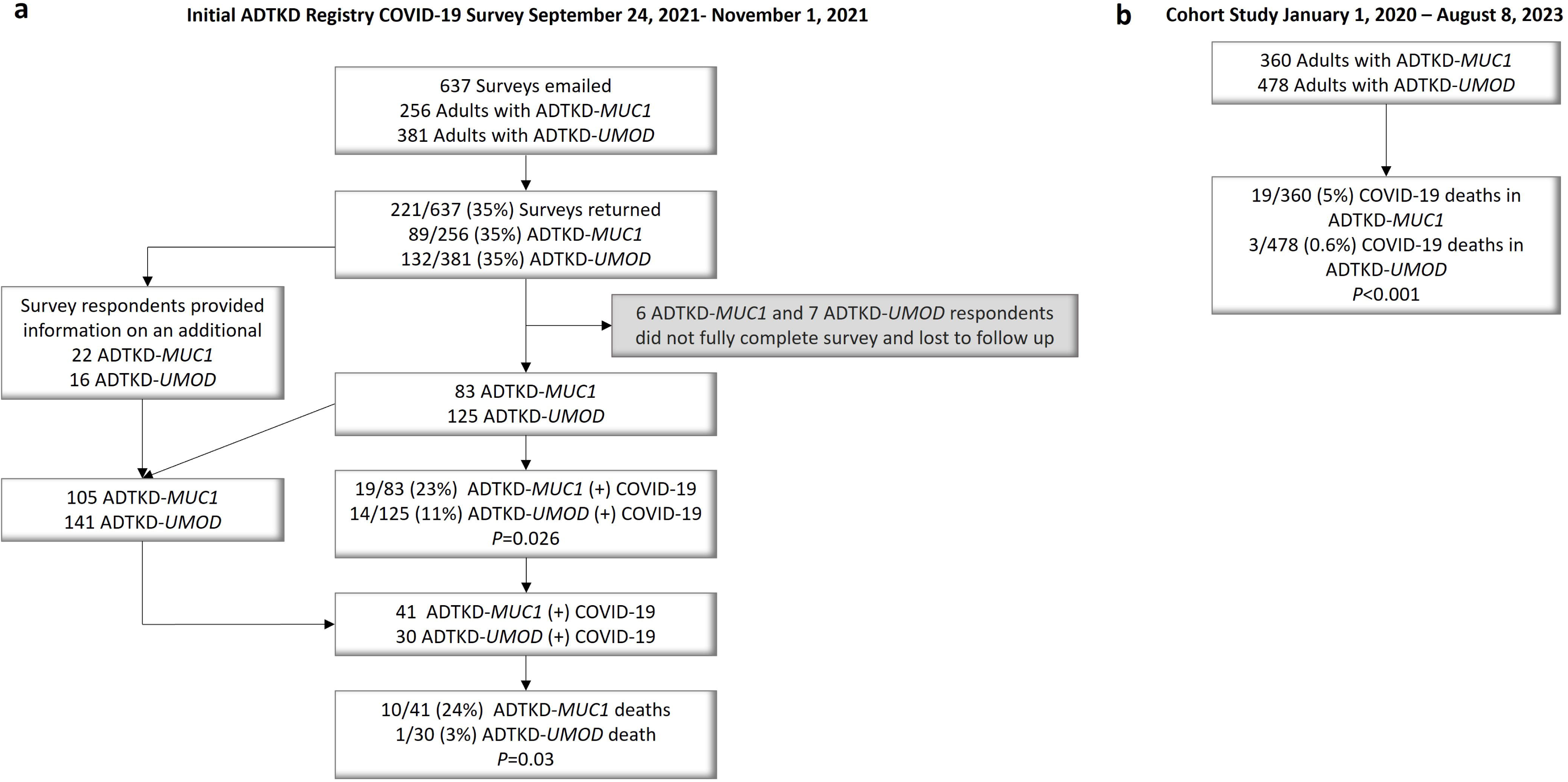
Flow diagram for the study. A) Initial ADTKD Registry COVID-19 Survey. B) Cohort study reviewing the ADTKD registry for COVID-19 related deaths.

Clinical characteristics were similar between the remaining 83 respondents with ADTKD-*MUC1* and 125 individuals with ADTKD-*UMOD* (see **Supplementary Table S2**). The respondents provided information on an additional 16 family members with ADTKD-*MUC1*, resulting in 105 individuals with ADTKD-*MUC1* and an additional nine family members with ADTKD-*UMOD*, resulting in 141 individuals (see **Figure 2a**).

### COVID-19 Outcomes During the Early Phase of the Pandemic

Wecompared COVID-19 outcomes in survey respondents with ADTKD-*MUC1* vs. ADTKD-*UMOD* in Sep 2021, after the Delta variant had peaked in the US (see **Figure 2a**). Of 83 ADTKD-*MUC1* individuals, 19 (22%) developed COVID-19 infection vs. 14/125 (11%) of ADTKD-*UMOD* individuals (odds ratio (OR) 2.35 (95%CI 1.60-3.11, *P* = 0.026). **Table 1** shows a comparison of the individuals who developed COVID-19 by disease type. There was no statistical difference between individuals with regards to age, gender, or body mass index, though numbers were small in each group. At the time of COVID-19 infection, 16% of ADTKD-*MUC1* and 21% of ADTKD-*UMOD* individuals were unvaccinated. ADTKD-*MUC1* individuals who did not develop COVID-19 were more likely to be vaccinated, but this is likely related to bias, as ADTKD-*MUC1* individuals who developed COVID-19 may have developed it several months prior to the survey and often did so before the vaccine was available, while individuals who did not develop COVID-19 could have received the vaccine up until the time of the survey. As stated above, in a univariate model the odds ratio for developing COVID-19 infection was 2.35 for ADTKD-*MUC1* vs. ADTKD-*UMOD*. Kidney transplantation status, body mass index, age, and gender were not significant in univariate models or when added to ADTKD type in a multivariate model.

**Table 1.**
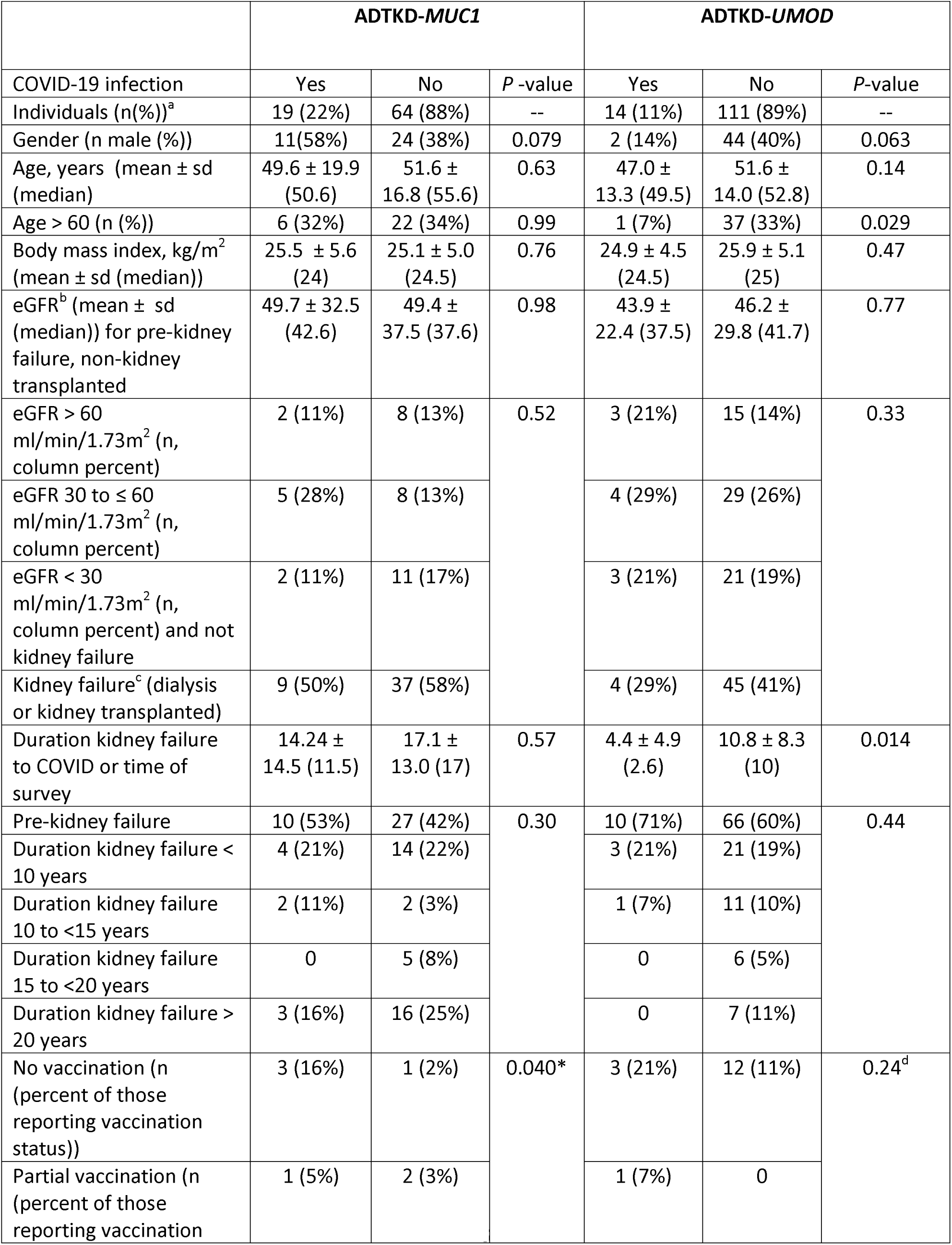
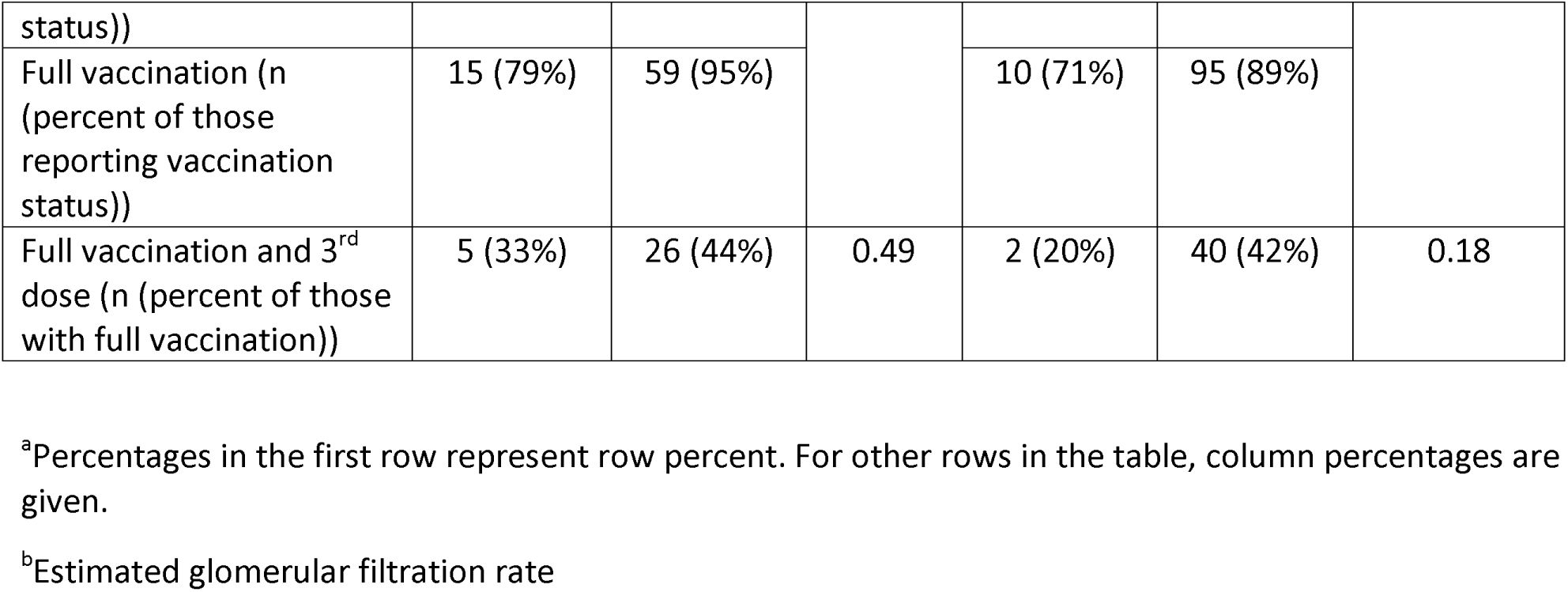
Characteristics of individuals developing COVID-19 by disease type.

At this stage of the analysis, we started to include data that respondents had provided regarding affected family members with COVID-19 (see **Figure 2a**), including 16 individuals with ADTKD-*MUC1* and nine individuals with ADTKD-*UMOD*, all of whom were previously in our registry. Of the total number of cases of COVID-19 identified, 10/41 (24%) of ADTKD-*MUC1* COVID-19 individuals died vs. 1/30 (3%) ADTKD *UMOD* COVID-19 individuals. Using a generalized estimating equation to account for the family structure, we found an odds ratio of mortality in ADTKD-*MUC1* vs. ADTKD-*UMOD* of 9.21, 95% CI 1.22-69.32, *P* = 0.03.

Thus, in the early and more virulent phase of the COVID-19 epidemic, patients with ADTKD-*MUC1* were more likely to develop COVID-19 and were much more likely to die from COVID-19 compared to patients with ADTKD-*UMOD*.

### International Assessment of Mortality in COVID-19 According to ADTKD Type

We then queried collaborators (CS, JAS, PJC, LR) from three other academic centers in November 2021 and found 3/24 (13%) deaths in individuals with ADTKD-*MUC1* who developed COVID-19 vs. 0 deaths in 11 individuals with ADTKD-*UMOD* who developed COVID-19. Six of the 24 individuals with ADTKD-*MUC1* were kidney transplanted vs 4/11 in the ADTKD-*UMOD* group. The mean age of the ADTKD-*MUC1* group was 50.8 ± 14.4 years vs. 53.7 ± 12.3 years in the ADTKD-*UMOD* group.

### Plasma Mucin-1 Levels in ADTKD-MUC1 in Infected vs. Uninfected ADTKD-*MUC1* Patients

To determine whether the increased infection rate was associated with decreased mucin-1 production, we compared plasma CA15-3 (mucin-1) levels obtained as part of a prior study^19^ on many of the survey individuals. The mean CA15-3 level for 14 COVD-19 infected ADTKD-*MUC1* individuals was 7.06±4.12 U/mL vs. 10.21±4.02 U/mL for 27 uninfected ADTKD-*MUC1* individuals (*P* = 0.035). For 11 COVID-19 infected ADTKD-*UMOD* individuals, the mean CA15-3 level was 14.18±2.35 U/mL vs 13.28±5.53 U/mL for 49 uninfected individuals (*P* = 0.60).

### Observational Cohort Study of COVID-19 Deaths in ADTKD-*MUC1*

We then continued an observational cohort study of COVID-19 deaths (see **Figure 1**, **Figure 2b**). **Table 2** shows the characteristics of adults in our registry according to ADTKD type. 19/360 (5%) individuals with ADTKD-*MUC1* died from COVID-19 vs. 3/478 (0.6%) individuals with ADTKD-*UMOD* (*P* = 0.0007). The 19 deaths in ADTKD-*MUC1* individuals occurred in 16 families. The three deaths in ADTKD-*UMOD* individuals occurred in three families. **Table 3** shows the characteristics of patients who died of COVID-19 according to ADTKD type. Univariate and multivariate logistic models were then created with death from COVID-19 as the binary outcome (see **Table 4**). Patients with ADTKD-*MUC1* had an eight-fold increased risk of death (8.4 (29-29.5%, *P* = 0.009)) vs. ADTKD-*UMOD* after adjustment for body mass index, kidney transplant status, and age. Of the patients with ADTKD-*MUC1* who died (see **Supplemental Table S3**), 84% had undergone kidney transplant vs. 53% in ADTKD-*MUC1* patients who had not died (*P* = 0.008). 43% of the ADTKD-*MUC1* patients who died had reached KF > 10 years prior to death vs. 26% of ADTKD-*MUC1* patients in our registry who did not die of ADTKD-*MUC1* (*P* = 0.001). **Table S3** shows a comparison of characteristics for patients with ADTKD-*MUC1* who died vs. those who did not.

**Table 2.** Characteristics ADTKD-*MUC1* and ADTKD-*UMOD* adults in registry.

| Characteristic | ADTKD-*MUC1* | ADTKD-*UMOD* | *P* -value<br>Odd Ratio (OR)<br>given with 95%<br>Confidence<br>Interval (CI) where<br>applicable | *P*-value<br>grouping for<br>Kidney Failure<br>Modality<br>(No Kidney<br>Failure,<br>Dialysis,<br>Kidney<br>Transplant)<br>To consider a<br>change to the<br>table |
| --- | --- | --- | --- | --- |
| n | 360 | 478 |  |  |
| Gender (% male) | 41% | 45% | 0.19 |  |
| Age, y (mean ± s.d.<br>(median)) | 47.40 ± 15.8<br>(47.74) | 47.38 ± 14.89<br>(48.07) | 0.97 |  |
| Age > 60 (n,%) | 86 (24%) | 118 (25%) | 0.68 |  |
| Race (% White) | 86% | 95% |  |  |
| BMI | 26.22 ± 5.58<br>(25.5) | 26.86 ± 5.75<br>(25.8) | 0.47 |  |
| BMI greater than 25<br>n(%) | 161 (55%) | 211 (56%) | 0.89 |  |
| On dialysis | 34 (9%) | 26 (5%) | 0.0027 | 0.0011 |
| Living with kidney<br>transplant | 166 (46%) | 164 (34%) | 0.0037 |  |
| Not on dialysis or kidney<br>transplanted (n, column<br>percent) | 159 (44%) | 287 (60%) | <0.0001 |  |
| Duration kidney failure<sup>a</sup><br>< 10 years | 104 (29%) | 110 (38%) |  |  |
| Duration kidney failure<br>10 to ≤ 15 years | 25 (7%) | 33 (11%) |  |  |
| Duration kidney failure<br>15 to ≤ 20 years | 31 (9%) | 18 (6%) |  |  |
| Duration kidney failure<br>> 20 years | 41 (11%) | 25 (9%) |  |  |

<sup>a</sup>Duration of kidney failure until the start of 2020

**Table 3.** Characteristics of individuals who died of COVID-19 related infection.

| Characteristic | ADTKD-*MUC1* | ADTKD-*UMOD* | *P*-value<br>Odd Ratio (OR) given<br>with 95% Confidence<br>Interval (CI) where<br>applicable |
| --- | --- | --- | --- |
| Individuals (n) | 19 | 3 |  |
| Gender (n male (%)) | 11 (58%) | 2 (67%) | 0.89 |
| Age, years (mean ± sd) | 56.49 ± 8.45 (57.62) | 71.44 ± 7.25 (74.80) | 0.0002 |
| Age > 60 years (n (%)) | 7 (37%) | 3 (100%) | < 0.0001<sup>b</sup> |
| Body mass index,<br>kg/m<sup>2</sup> (mean ± sd) | 30.78 ± 6.46 (30.74) | 25.56 ± 2.02 (25.11) | 0.02 |
| Kidney transplanted (n<br>(%)) | 16 (84%) | 3 (100%) | 0.46<sup>b</sup> |
| Pre-kidney failure (n<br>(%)) | 2 (11%) | 0 | 0.01<sup>b</sup> |
| No vaccination (n<br>(percent of those<br>reporting vaccination<br>status)) | 11/16 (69%) | 1 (33%) | 0.23 |
| Average length of<br>hospital stay (mean ±<br>sd) | 18.67 ± 10.80 (21) | 12 ± 5 (12) | 0.15 |
| Required mechanical<br>ventilation | 18/18 (100%) | 2 (67%) | 0.01<sup>b</sup> |
| Years of kidney failure<sup>a</sup> | 12.86 ± 10.84 (9) | 21.93 ± 14.96 (13.76) | 0.45 |
| >10 years kidney failure<sup>a</sup> | 9/17 (53%) | 3 (100%) | 0.003<sup>b</sup> |

<sup>a</sup>Years of kidney failure are years from start of kidney failure until contracting COVID-19.
<sup>b</sup> *P*-values calculated using permutation test.

**Table 4.** Univariate and multivariate logistic regression models with death from COVID-19 as the outcome variable.

|  |  |  | Univariate |  | Multivariate |  |
| --- | --- | --- | --- | --- | --- | --- |
| Parameter | Reference | Risk group | Odds Ratio | P-value | Odds Ratio | P-value |
| ADTKD | *UMOD* | *MUC1* | 8.54 (2.49-29.36) | 0.0007 | 8.38 (2.86-29.47) | 0.0009 |
| Body mass index (kg/m<sup>2</sup>) |  | Increase by 1 kg/m<sup>2</sup> | 1.09 (1.03-1.15) | 0.003 | 1.11 (1.04-1.18) | 0.002 |
| Kidney Transplant Status | No | Yes | 9.66 (2.77-33.67) | 0.0003 | 5.49 (1.58-19.05) | 0.007 |
| Age (years) |  | Increase by 1 year | 1.06 (1.03-1.08) | <0.0001 | 1.04 (1.01-1.07) | 0.003 |

## DISCUSSION

This investigation found that patients with ADTKD-*MUC1* are at a markedly increased risk of death from COVID-19, with 5% of adult ADTKD-*MUC1* patients dying of COVID-19 during the three years of the pandemic vs. 0.6% of ADTKD-*UMOD* adults (*P* = 0.0007). Most, but not all, of the deaths occurred in patients who had undergone kidney transplantation, though the death rate was not similarly high in transplanted patients with ADTKD-*UMOD*. Patients died of respiratory complications, and many had prolonged hospitalizations. While most of the deaths occurred early in the pandemic, deaths have continued to occur.

While patient numbers were small due to the rarity of ADTKD, we showed consistent results with several different analyses: (1) We identified a 2.35 (95% confidence interval 1.60-3.11) increased odds of COVID-19 infection in ADTKD-*MUC1* individuals early in the pandemic, with 23% of ADTKD-*MUC1* individuals developing COVID-19 vs. 11% of ADTKD-*UMOD* individuals (*P* = 0.026). (2) There was an early increased risk of death, with 24% of individuals with ADTKD-*MUC1* dying from COVID-19 vs. 3% in individuals with ADTKD-*UMOD* (*P* = 0.03). (3) We found that the mean steady state CA15-3 level at least 30 days prior to COVID-19 infection^19^ for 14 COVD-19 infected ADTKD-*MUC1* individuals was 7.06 ± 4.12 U/mL vs. 10.21 ± 4.02 U/mL for 27 uninfected individuals (*P* = 0.035). (4) Our longitudinal study then revealed an 8-fold risk of death from COVID-19 in patients with ADTKD-*MUC1* vs. ADTKD-*UMOD* (*P* = 0.0009). Thus, compared to ADTKD-*UMOD* individuals, ADTKD-*MUC1* individuals had a higher COVID-19 infection rate and were more likely to die of COVID-19. ADTKD-*MUC1* individuals were more likely to contract COVID-19 if their plasma CA15-3 levels were low.

In the early stages of COVID-19, eight (36%) of twenty-two ADTKD-*MUC1* kidney transplanted patients died, compared to one (25%) of four ADTKD-*UMOD* kidney transplanted patients. In a meta-analysis of COVID-19 outcomes in patients with kidney transplants during the Delta strain of SARS-CoV-2^25^, the mortality rate was approximately 21%. While it is difficult to compare patient characteristics between studies, ADTKD-*MUC1* patients are in general quite healthy with no comorbidities except kidney disease. For example, in the meta-analysis^25^, there was an approximate 10% prevalence of diabetes and 8% prevalence of cardiovascular disease.

There were several weaknesses in our study. Most importantly, the sample size was small. Fortunately, we have the largest registry of ADTKD, with over 280 families and 800 individuals, and the response rate to our survey was good. However, by definition, any study of rare diseases will be associated with a small patient population. Another potential weakness was ascertainment and selection bias. We attempted to eliminate as many sources of bias as possible. Our registry has been historically designed to study all individuals with ADTKD-*UMOD* and ADTKD-*MUC1* in a similar manner. The surveys were sent out in an un-biased fashion, and the characteristics of respondents was similar for ADTKD-*UMOD* and ADKTD-*MUC1*. However, there could be unanticipated forms of bias that we did not identify. Another weakness of our study is that we did not examine the prevalence of other respiratory infections according to disease type. We plan to do this in the future.

The results of our investigation provide unique clinical information regarding the role of mucin-1 in COVID-19 infection. Our investigation shows that ADTKD-*MUC1* individuals are at increased risk from COVID-19, based on lower mucin-1 levels prior to infection. There is emerging information that mucin-1 in the nares and lungs serves as an initial protective barrier against infection. Lai et al. showed that mucin-1 inhibited SARS-CoV-2 viral attachment, entry, and post-entry replication.^16^ Biering et al. performed genome-wide bidirectional CRISPR screens to define host-pathogen interactions required for facilitating or restricting SARS-CoV-2 in a human lung cell line and identified mucin-1 as an important host defense factor.^14^ The investigators then showed an antiviral role for membrane-anchored mucins (including mucin-1) *in vitro* and in mouse models of SARS-CoV-2 infection. Membrane-bound mucins specifically inhibited spike-mediated viral entry into lung cells. Chatterjee et al. showed that mucin-1 is highly expressed on the ACE2-positive respiratory epithelial cells, and that removal of the mucin domains enhances spike binding^17^. Our investigation provides complementary evidence in humans. Patients with ADTKD-*MUC1* have mutations that prevent expression of the mucin domain, and these patients are at markedly increased risk of infectin and death from COVID-19.

While there is evidence that decreased production of mucin-1 is associated with an increased risk from COVID-19, there is also evidence that increased production of mucin-1 may be harmful. Mucin-1 participates in the cytokine storm that occurs with COVID-19 and leads to an inflammatory response that is highly damaging. In studies of individuals who do not have ADTKD-*MUC1*, elevated mucin-1 content of bronchoalveolar fluid^26^, airway mucus^27^, and blood^28–31^ at the time of infection is associated with worse and more severe COVID-19 outcomes. A recent genome-wide association study ^32^ identified the rs41264915 intronic variant *THBS3* (associated with increased mucin-1 expression) as being associated with critical COVID-19. Another GWAS study suggested that increased mucin-1 expression is associated with more severe COVID-19, while other GWAS studies have not found this association.^33^ Thus, lowering mucin-1 production during infection has been postulated as a potential therapy for COVID-19^34^.

Compounds increasing mucin-1 production could be prophylactic for individuals with low mucin-1 levels, whereas compounds reducing mucin-1 production could be therapeutic during acute infection. The finding that both pathologically low and elevated clinical characteristics are associated with increased mortality is well known and has also been noted with body mass index^35^, sleep time^36^, and many laboratory parameters^37^.

Given the increased risk associated with ADTKD-*MUC1* individuals from COVID-19, it would be prudent for families with an unknown cause of ADTKD (chronic kidney disease, autosomal dominant transmission, bland urinary sediment) to be tested for a *MUC1* mutation. Unfortunately, genotyping for the vast majority of *MUC1* mutations is currently not performed in any commercial genetics laboratory, as these mutations are not identified by standard Sanger sequencing.^38^ Mucin-1 plays an important role in other respiratory tract infections^14^, and we need to extend our studies in patients with ADTKD-*MUC1* to other pulmonary infections.

In summary, this investigation shows that individuals with ADTKD-*MUC1* are at increased risk of COVID-19 infection and mortality, especially for immunosuppressed and kidney transplanted patients exposed to more virulent COVID-19 variants. ADTKD-*MUC1* patients with low plasma mucin-1 levels may be especially at increased risk. We be lieve that clinicians should consider more aggressive preventive care and treatment for COVID-19 in patients with ADTKD-*MUC1*.

## Supporting information

Supplementary Material covid-19 adtkd

## Data Availability

All data produced in the present study are available upon reasonable request to the authors

## DATA SHARING

We are very happy to work with any groups interested in studying this condition and providing genetic information that can satisfy their research requests and protect patient confidentiality. An anonymized dataset analyzed in the study will be available from the European Genome-Phenome Archive (EGA-archive.org), with request of data access.

## ACKNOWLEDGEMENTS

SK and colleagues were supported by the OP Integrated Infrastructure, the project: Research on COVID-19 progressive diagnostic methods and biomarkers useful in early detection of individuals at increased risk of severe disease, ITMS: 313011ATA2, co-financed by the European Regional Development Fund and by the Slovak Research and Development Agency under the Contract no. PP-COVID-20-0056, by the Ministry of Health of the Czech Republic (grants NU21-07-00033, NU22-A-123), the Ministry of Education of the Czech Republic (grant LTAUSA19068) and by institutional programs of Charles University in Prague (UNCE/MED/007). JAS and HRM are funded by the Medical Research Council. The National Center for Medical Genomics (LM2023067) kindly provided sequencing and genotyping. AJB was funded by NIH-NIDDK R21 DK106584, CKD Biomarkers Consortium Pilot and Feasibility Studies Program funded by the NIH-NIDDK (U01 DK103225), the Slim Health Foundation, the Black-Brogan Foundation, Soli Deo Gloria. EE reports funds from the Royal College of Surgeons in Ireland StAR PhD. CD received funding from The Slim Health Foundation and the European Union’s Horizon 2020 Program, the Government of Cyprus and the University of Cyprus, under grant agreement number 857122. The authors thank all participating patients and families who participated in this study.

## CONTRIBUTORS

Conceptualization: AJB; Data curation: KOK, AT, CS, GP, HRM, REH; Formal analysis: AJB, KOK, AHW; Funding acquisition: AJB, SK; Investigation: KOK, AT, LM, VR, APR-C, JAS, EO, HRM, GP, CD, CS, PJC, REH, EE, PV, KH, MZ; Methodology: AJB, KOK, DS, AV, LP, PV, KH, MZ, SK; Project Administration: KOK, AT; Resources: KOK, AHW, AT, LM, VR, JAS, EO, HRM, GP, CD, CS, PJC, REH, EE, CI, MP, MR, PV, KH, MZ, SK, AJB; Software: KOK, AHW, AT; Supervision: AJB, SK, TZ, CS, PJC, JAS; Validation: KOK, AHW; Visualization: KOK, AHW; Writing-original draft: AJB, KOK, AHW; Writing-review & editing: KOK, AHW, AT, LM, VR, JAS, EO, HRM, GP, CD, CS, PJC, REH, EE, CI, PV, KH, MZ, SK, and AJB.

## LIST OF SUPPLEMENTARY MATERIALS

Supplementary Table S1.

Supplementary Table S2.

Supplementary Table S3.

Supplementary Survey.

