## Supplementary Material covid-19 adtkd for "Eight-Fold Increased COVID-19 Mortality in Autosomal Dominant Tubulointerstitial Kidney Disease due to *MUC1* Mutations: An Observational Study"

This supplementary material has been provided by the authors to give readers additional information about their work.

#### TABLE OF CONTENTS

**Supplementary Table S1** (p. 2)

Characteristics of genetically affected individuals who were respondents and non-respondents to the survey.

**Supplementary Table S2** (p.3-4)

Characteristics ADTKD-*MUC1* and ADTKD-*UMOD* survey respondents.

**Supplementary Table S3** (p. 5)

Characteristics ADTKD-*MUC1* patients who survived and died of COVID-19.

**Supplementary Survey** (p. 6-16)

Provided is the PDF generated by REDCap of the COVID-19 Effects in Inherited Kidney Disease Survey. The survey was built to capture information about the individual, their personal experience with COVID19 and information about affected family members who experienced COVID-19. Based upon the number of family members who experienced COVID-19 entered, separate “COVID-19 Family Information” forms populated for the individual to complete.

**Supplementary Table S1.** Characteristics of genetically affected individuals who were respondents and non-respondents to the survey.

| Characteristic | Respondents | Non-Respondents | *P* value |
|---|---|---|---|
| n | 221 | 416 | |
| Gender (% male) | 38.9% | 45.4% | 0.12 |
| Age mean +/- SD (median) | 50.19±15.19 (52) | 47.02±15.99 (47) | 0.018 |
| Race (% White) | 96.8% | 89.7% | 0.02 |
| %of ADTKD-*MUC1* individuals | 40.3% (row 34.8%) | 40.1% (row 65.2%) | 0.94 |
| % of ADTKD-*UMOD* individuals | 59.7% (row 34.6%) | 59.9% (row 65.4%) | |
| eGFR > 60 ml/min/1.73m$^2$ (n, column percent) | 31 (14.2%) | 77 (19.3%) | 0.012 |
| eGFR 30 to ≤ 60 ml/min/1.73m$^2$ (n, column percent) | 50 (22.8%) | 86 (21.6%) | |
| eGFR < 30 ml/min/1.73m$^2$ and not esrd (n, column percent) | 37 (16.9%) | 57 (14.3%) | |
| Kidney Failure, Kidney transplanted (n, column percent) | 94 (42.9%) | 95 (23.9%) | |
| Kidney Failure, non-kidney transplanted on dialysis (n, column percent) | 7 (3.2%) | 83 (20.9%) | |
| No Kidney Failure | 120 (54.3%) | 231 (56.6%) | 0.62 |
| Duration Kidney Failure < 10 Years | 44 (19.9%) | 77 (18.9%) | |
| Duration Kidney failure 10-15 Years | 17 (7.7%) | 31 (7.6%) | |
| Duration Kidney Failure 15-20 Years | 12 (5.4%) | 20 (4.9%) | |
| Duration Kidney Failure > 20 Years | 28 (12.7%) | 49 (12.0%) | |

**Supplementary Table S2.** Characteristics ADTKD-*MUC1* and ADTKD-*UMOD* survey respondents

| Characteristic | ADTKD-*MUC1* | ADTKD-*UMOD* | *P*-value Odd Ratio (OR) given with 95% Confidence Interval (CI) where applicable |
|---|---|---|---|
| N | 83 | 125 | |
| Gender (% male) | 42% | 37% | 0.42 |
| Age, y (mean ± s.d. (median)) | 51.13 ± 17.45 (55.5) | 51.07 ± 13.93 (52.3) | 0.97 |
| Race (% White) | 95% | 98% | 0.34 |
| eGFR[a] for non-transplanted, non-dialysis (mean ± s.d. (median)) [b] | 49.47 ± 35.86 (41.1) | 45.93 ± 28.79 (40) | 0.60 |
| eGFR > 60 ml/min/1.73m$^2$ (N ( column percent)) [b] | 10 (12%) | 18 (15%) | |
| eGFR 30 to ≤ 60 ml/min/1.73m$^2$ (N (column percent)) | 13 (16%) | 33 (27%) | |
| eGFR < 30 ml/min/1.73m$^2$ (N (column percent)) | 13 (16%) | 24 (19%) | 0.041 |
| Status post kidney transplant (N (column percent)) | 45 (55%) | 48 (39%) | |
| Receiving dialysis (N (column percent)) | 1 (1%) | 1 (1%) | |
| Not on dialysis or kidney transplanted (N (column percent)) | 37 (45%) | 76 (61%) | |
| Duration Kidney Failure < 10 years | 18 (22%) | 24 (19%) | |
| Duration Kidney Failure 10 to ≤ 15 years | 4 (5%) | 12 (10%) | 0.0039 |
| Duration Kidney Failure 15 to ≤ 20 years | 5 (6%) | 6 (5%) | |
| Duration Kidney Failure > 20 years | 19 (23%) | 7 (6%) | |
| Reported vaccination status (N) | 82 | 125 | |
| No vaccination (N (percent of those reporting vaccination status)) | 4 (5%) | 15 (12%) | 0.34 |
| Partial vaccination (N (percent of those reporting vaccination status)) | 3 (4%) | 1 (1%) | |

| | | | |
|---|---|---|---|
| Full vaccination (N (percent of those reporting vaccination status)) | 74 (90%) | 105 (84%) | |
| Full vaccination and 3rd dose (N (percent of those with full vaccination)) | 31 (42%) | 42 (40%) | 0.82 |
| Developed COVID-19 (N (% of individuals)) | 19 (23%) | 14 (11%) | 0.026 OR=2.35 (95%CI 1.60-3.11) |

[a]Estimated glomerular filtration rate

[b]The eGFR measurements were not available on one individual with ADTKD-*MUC1* and one individual with ADTKD-*UMOD*.

**Supplementary Table S3**. Characteristics ADTKD-*MUC1* patients who survived and died of COVID-19

<table>
<tr><th>Characteristic</th><th>ADTKD-MUC1<br>Controls</th><th>ADTKD-MUC1<br>Deaths</th><th>P-value<br>Odd Ratio (OR) given with 95% Confidence Interval (CI) where applicable</th></tr>
<tr><td>n</td><td>341</td><td>19</td><td></td></tr>
<tr><td>Gender (% male)</td><td>202 (59%)</td><td>11 (58%)</td><td>0.87</td></tr>
<tr><td>Age, y (mean ± s.d. (median))</td><td>46.89 ± 15.97 (46.62)</td><td>56.98 ± 8.45 (57.62)</td><td>< 0.0001</td></tr>
<tr><td>Age > 60 (%)</td><td>79 (23%)</td><td>7 (37%)</td><td>0.18</td></tr>
<tr><td>Race (% White)</td><td>292 (86%)</td><td>18 (95%)</td><td>0.29</td></tr>
<tr><td>BMI</td><td>25.93 ± 5.40 (25.36)</td><td>30.78 ± 6.46 (30.74)</td><td>0.004</td></tr>
<tr><td>BMI > 25 kg/m$^2$ (%)</td><td>146 (53%)</td><td>15 (83%)</td><td>0.02<br>4.15 (1.22-14.06)</td></tr>
<tr><td>On dialysis</td><td>33 (10%)</td><td>1 (5%)</td><td>0.52</td></tr>
<tr><td>Living with transplant</td><td>150 (44%)</td><td>16 (84%)</td><td>0.008<br>5.89 (1.70-20.42)</td></tr>
<tr><td>Not on dialysis or transplanted (n, column percent)</td><td>157 (46%)</td><td>2 (11%)</td><td rowspan="5">0.001</td></tr>
<tr><td>Duration Kidney Failure[c] < 10 years</td><td>95 (27%)</td><td>9 (47%)</td></tr>
<tr><td>Duration Kidney Failure 10 to ≤ 15 years</td><td>23 (7%)</td><td>2 (11%)</td></tr>
<tr><td>Duration Kidney Failure 15 to ≤ 20 years</td><td>28 (8%)</td><td>3 (16%)</td></tr>
<tr><td>Duration Kidney Failure > 20 years</td><td>38 (11%)</td><td>3 (16%)</td></tr>
</table>

### COVID-19 and ADTKD Consent Form

Dear Study Participant,

We are conducting a survey called “COVID-19 in Inherited Kidney Disease.” The results will be incredibly helpful to us in the future as we work to improve healthcare of patients with inherited kidney disease.

You will be asked to read a consent form first, if you agree, you will continue to the survey questions. If you do not agree, you will not be asked any further questions.

If you have any questions, please contact.

Thank you,

Wake Forest School of Medicine Rare Inherited Kidney Disease Research Team

SUBSTUDY: COVID 19 IN INHERITED KIDNEY DISEASE SURVEY
Informed Consent Form to Participate in Research
Anthony Bleyer, MD, MS Principal Investigator SUMMARY
You are invited to participate in a research study. The purpose of this research is to find out the effects of COVID-19 in the families we study and to see if families are at an increased risk of COVID-19. You are invited to be in this study because your family has inherited kidney disease. Your participation in this research will involve completion of an online survey that will take 20-30 minutes to complete.

Participation in this study will involve an online survey. All research studies involve some risks. A risk to this study that you should be aware of is a slight risk of a breach of confidentiality. You may not benefit from participation in this study.

Your participation in this study is voluntary. You do not have to participate in this study if you do not want to. There may be other choices available to you. Some other choices may include not participating in the COVID-19 survey. You will not lose any services, benefits, or rights you would normally have if you choose not to participate.

The remainder of this form contains a more complete description of this study. Please read this description carefully. You can ask any questions if you need help deciding whether to join the study. The person in charge of this study is Anthony Bleyer, MD . If you have questions, suggestions, or concerns regarding this study or you want to withdraw from the study his contact information is: 336-716-4650 or email at. If you have any questions, suggestions or concerns about your rights as a volunteer in this research, contact the Institutional Review Board at 336-716-4542 or the Research Subject Advocate at Wake Forest at 336-716-8372.

HOW MANY PEOPLE WILL TAKE PART IN THE STUDY? Approximately 1300 people will be asked to take part in this study.

WHAT ARE THE RISKS OF THE STUDY? There is a slight risk of a breach of confidentiality. We will do our best to protect your confidential information. Efforts, such as coding research records, keeping research records secure and allowing only authorized people to have access to research records, will be made to keep your information safe.

ARE THERE BENEFITS TO TAKING PART IN THE STUDY? You are not expected to receive any direct benefit from taking part in this research study. We hope the information learned from this study will benefit other people in the future.

WHAT ARE THE COSTS? All study costs, related directly to the study, will be paid for by the study. Costs for your regular medical care, which are not related to this study, will be your own responsibility.

WILL YOUR RESEARCH RECORDS BE CONFIDENTIAL? The results of this research study may be presented at scientific or medical meetings or published in scientific journals. Your identity and/or your personal health information will not be disclosed unless it is authorized by you, required by law, or necessary to protect the safety of yourself or others. There is always some risk that even de-identified information might be re-identified.

Participant information may be provided to Federal and other regulatory agencies as required. The Food and Drug Administration (FDA), for example, may inspect research records and learn your identity if this study falls within its jurisdiction.

WILL YOU BE PAID FOR PARTICIPATING?

You will receive no payment or other compensation for taking part in this study.

What About My Health Information? In this research study, any new information we collect from you about your health or behaviors is considered Protected Health Information. The information we will collect for this research study includes: Name and Date of Birth.

We will make every effort to keep your Protected Health Information private. We will store records of your Protected Health Information in a cabinet in a locked office or on a password protected computer.

Your personal health information and information that identifies you ("your health information") may be given to others during and after the study. This is for reasons such as to carry out the study, to determine the results of the study, to make sure the study is being done correctly.

Some of the people, agencies and businesses that may receive and use your health information are the research sponsor; the Institutional Review Board; representatives of Wake Forest University Health Sciences and North Carolina Baptist Hospital; representatives from government agencies such as the Food and Drug Administration (FDA) or the Office of Human Research Protections (OHRP), the Department of Health and Human Services (DHHS) and similar agencies in other countries.

Some of these people, agencies and businesses may further disclose your health information. If disclosed by them, your health information may no longer be covered by federal or state privacy regulations. Your health information may be disclosed if required by law. Your health information may be used to create information that does not directly identify you. This information may be used by other researchers. You will not be directly identified in any publication or presentation that may result from this study unless there are photographs or recorded media which are identifiable.

If required by law or court order, we might also have to share your Protected Health Information with a judge, law enforcement officer, government agencies, or others. If your Protected Health Information is shared with any of these groups it may no longer be protected by federal or state privacy rules.

Any Protected Health Information collected from you in this study that is maintained in the research records will be kept for at least six years after the study is finished. At that time any research information not already in your medical record will either be destroyed or it will be de-identified You will not be able to obtain a copy of your Protected Health Information in the research records until all activities in the study are completely finished.

You can tell Dr. Anthony Bleyer that you want to take away your permission to use and share your Protected Health Information at any time by sending a letter to this address: Anthony Bleyer, MD, MS Wake Forest School of Medicine
Section on Nephrology
Medical Center Blvd
Winston-Salem, NC 27157  However, if you take away permission to use your Protected Health Information you will not be able to be in the study any longer. We will stop collecting any more information about you, but any information we have already collected can still be used for the purposes of the research study.

WHAT ARE MY RIGHTS AS A RESEARCH STUDY PARTICIPANT?

You may choose not to take part or you may leave the study at any time. If you decide to stop participating in the study we encourage you to talk to the investigators or study staff. The investigators also have the right to stop your participation in the study at any time. This could be because new information becomes available. Information about you may be removed from the study data and could be used for future research or shared with other researchers without additional consent from you.

By continuing, I agree to take part in this study. I authorize the use and disclosure of my health information as described in this consent and authorization form. I have had a chance to ask questions about being in this study and have those questions answered. By taking part in the study, I am not releasing or agreeing to release the investigator, the sponsor, the institution or its agents from liability for negligence.

PLEASE SELECT ONE OF THE FOLLOWING:

◯ YES, I consent to participate in this study
◯ NO, I do NOT consent to participate in this study

### COVID19 and ADTKD Survey

Thank you for participating in our COVID-19 and ADTKD study.

Please note this is a self-building survey. These first 4 required questions are used to build your survey.

Based on your answers, you may be asked additional questions, and could take up to 30 minutes to complete. If needed, there are options to save and return later to complete the survey.

#### Personal Health Information

Name: ______________________________

Date of Birth: ______________________________
(Month-Day-Year)

Do you have a known UMOD, MUC1, or REN mutation?

◯ Yes
◯ No
◯ Unknown

Your survey will be 2 parts. If you have experienced a COVID-19 infection, your vaccination status, and current kidney health.
Individual family members who are genetically affected and have kidney disease who have experienced a COVID-19 infection.

#### COVID-19 Information about Additional Family Members Who are Genetically Affected with the kidney disease that runs in your family

This next question asks about the number of family members with kidney disease that have had COVID-19. This includes family members with kidney disease who may have died from COVID-19.

Based on the number selected 0-10, you will have a short survey to complete for each family member.

For example, if 3 family members with kidney disease have had COVID-19, there will be 3 surveys, one for each family member. Names and DOB will not be asked for your family members.

If you have more than 10 family members who experienced COVID-19 infection, please contact

How many family members genetically affected with the kidney disease that runs in your family have had COVID besides you? This includes family members with kidney disease who may have died of COVID-19.

◯ 0
◯ 1
◯ 2
◯ 3
◯ 4
◯ 5
◯ 6
◯ 7
◯ 8
◯ 9
◯ 10

If at any time, you need to exit out of the survey prior to the completion, please see the following following instructions for your return options:

1. The top right corner, you will see the icon button labeled "Get link to my survey queue" - Select it
2. A window will appear (picture shown below) with two options: copy and paste the survey queue link or send the survey queue link in an email.
3. We suggest sending the return link to your e-mail. You will enter your email address and select send. REDCap will send you the return link.

If you have any questions or issues, please contact

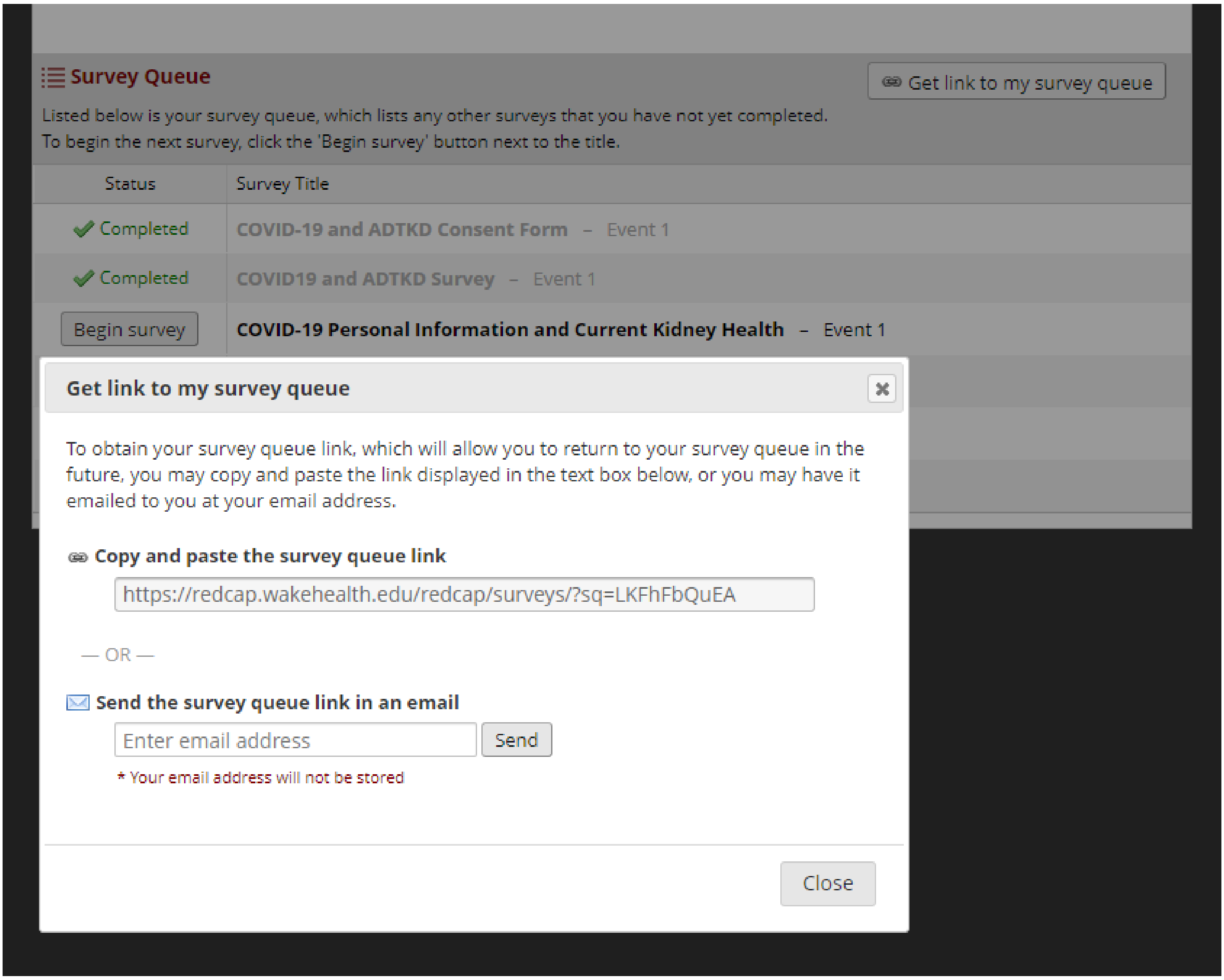

### COVID-19 Personal Information and Current Kidney Health

#### Personal COVID-19 Health Information

| | |
|---|---|
| Have you had COVID-19 that was confirmed by lab testing? | ◯ Yes ◯ No |
| Date it started: | ________________ (Month-Day-Year) |
| Were you hospitalized for COVID? | ◯ Yes ◯ No |
| How many days in the hospital? | ________________ |
| Were you in the ICU? | ◯ Yes ◯ No |
| Did you need a breathing tube? | ◯ Yes ◯ No |
| Did you receive treatment with a monoclonal antibody like remdesivir or toculizimab? | ◯ Yes ◯ No |
| Are there any things you would like to tell us about this event? | |
| Have you received the COVID vaccine? | ◯ Yes ◯ No |
| Which vaccine did you receive? | ◯ Pfizer<br>◯ Moderna<br>◯ Johnson & Johnson<br>◯ Other |
| What dates have you received a COVID vaccine? | |
| Date | ________________ |
| Date | ________________ |
| Date | ________________ |
| Other Name | ________________ |
| Date | ________________ |

| | |
|---|---|
| Date | __________________________________ |
| Date | __________________________________ |
| Are you currently on dialysis? | ◯ Yes ◯ No<br>(This answer is needed to populate other questions) |
| Type | ◯ Hemodialysis ◯ Peritoneal dialysis |
| What year did you start? | __________________________________ |
| Do you have a kidney transplant? | ◯ Yes ◯ No<br>(This answer is needed to populate other questions) |
| Year of kidney transplant? | __________________________________ |
| Are you currently being treated for cancer? | ◯ Yes ◯ No |
| Type: | __________________________________ |

#### Follow Up Regarding Your Current Kidney Health

Please list all of your serum creatinine (SCr) values from January 2020 until present.

Please list values even if you have received a transplant.

If you are a part of our study and have already submitted these to us, you do not need to fill out.

There are spaces for 8 SCr dates and values, there is a notes box to list additional SCr, or there is the option to upload a file that contains your values - please let know if you have additional information or have other questions.

If you do not have your serum creatinine values it is OK - we still would like you to fill out the rest of the survey.

| | |
|---|---|
| I have already shared my SCr values with Wake Forest: | ◯ Yes<br>(If no, please enter values, upload file, or continue to the next section of the survey. Thank you!) |

Thank you! We appreciate your participation in our study and working with us to update your Serum Creatinine values. You do not need to re-enter your values here.

| | |
|---|---|
| Date #1 | __________________________________ |
| Serum Creatinine #1 | __________________________________ |
| Date #2 | __________________________________ |

Serum Creatinine #2 __________________________________

Date #3 __________________________________

Serum Creatinine #3 __________________________________

Date #4 __________________________________

Serum Creatinine #4 __________________________________

Date #5 __________________________________

Serum Creatinine #5 __________________________________

Date #6 __________________________________

Serum Creatinine #6 __________________________________

Date #7 __________________________________

Serum Creatinine #7 __________________________________

Date #8 __________________________________

Serum Creatinine #8 __________________________________

I have Serum Creatinine values in a file to upload:

(upload file containing Scr values here)

Please list additional Serum Creatinines with date and value:

Thank you for your participation and help with this study for families with inherited kidney disease! No further information is needed at this time.

If you have any questions, please reach out to

| | |
|---|---|
| Is this information complete? Only select "YES" once you have entered in all of your information and feel like you have no more edits to make. | ◯ YES |

### COVID-19 Family Information

| | |
|---|---|
| Relation of the family member to yourself: | ◯ Mother ◯ Father ◯ Brother<br>◯ Sister ◯ Uncle ◯ Aunt<br>◯ Son ◯ Daughter ◯ Nephew<br>◯ Niece ◯ Cousin ◯ Grandmother<br>◯ Grandfather |
| What is the approximate date that COVID occurred? | ______________________<br>(If you do not know the exact date, please approximate month and year, example March 2021, use 03-01-2021) |
| The individual's approximate age when they developed COVID-19 | ______________________ |
| At the time of the COVID infection, was the person:<br><br>◯ Fully vaccinated<br>◯ Partially vaccinated<br>◯ Not vaccinated<br>◯ Unsure | |
| Did the person have a kidney transplant? | ◯ Yes ◯ No |
| Year? | ______________________ |
| Did the person have cancer that was being treated? | ◯ Yes ◯ No |
| Type | ______________________ |
| Was the person hospitalized for COVID? | ◯ Yes ◯ No |
| Approximately how many days in the hospital? | ______________________ |
| Was the person in the ICU? | ◯ Yes ◯ No |
| Did the person need a breathing tube? | ◯ Yes ◯ No |
| Did they receive treatment with a monoclonal antibody like remdesivir? | ◯ Yes ◯ No |
| Did the person die of COVID? | ◯ Yes ◯ No |
| Are there any things you would like to tell us about this event? | |

| | |
|---|---|
| Is this information complete? Only select "YES" once you have entered in all of your information and feel like you have no more edits to make. | ◯ YES |
